# The Relationship Between Suspected Adverse Drug Reactions Of HMG-Coa Reductase Inhibitors And Polypharmacology Using A National Registry Approach

**DOI:** 10.1101/2024.04.02.24305224

**Authors:** Hasan Yousaf, Alan M. Jones

## Abstract

**Aims:** The aim of this study was to explore the suspected adverse drug reaction (ADR) data of five licensed statins in the UK: atorvastatin, fluvastatin, pravastatin, rosuvastatin and simvastatin. A secondary aim was to determine if there was a link between the polypharmacological properties of the statins and their associated muscle-related side effects.

**Methods:** The chemical database of bioactive molecules with drug-like properties, European Molecular Biology Laboratory (ChEMBL) was used to obtain data on the pharmacological interactions of statins with human proteins. The Medicines and Healthcare products Regulatory Agency’s (MHRA) Yellow Card Scheme was used to obtain reports of suspected ADRs from 2018 to 2022. The OpenPrescribing database was used to obtain the prescribing rates for statistical interpretation.

**Results:** The study found no significant difference between the statins in causing ADRs across all organ classes (*X*^*2*^, *P* > .05). Fluvastatin was found to have a higher incidence of ADRs/100,000 *R*_*x*_ across multiple organ classes.

**Conclusion:** No significant difference was found between the suspected ADR incidence of the statins across all organ classes. No evidence of higher intensity statins causing more muscle symptoms than moderate intensity statins was found.

## Introduction

Statins are a group of lipid-lowering drugs that act by inhibiting 3-hydroxy-3-methylglutaryl (HMG)-coenzyme A reductase, the rate-limiting enzyme for the synthesis of mevalonic acid from HMG-coenzyme A.[1] Mevalonic acid is converted *via* precursor molecules into cholesterol. By inhibiting cholesterol synthesis, the body upregulates low-density lipoprotein (LDL) receptors causing a decrease in plasma LDL-cholesterol.[2] Elevated LDL-cholesterol is associated with an increased risk of myocardial infarction (MI) and atherosclerotic cardiovascular disease (CVD), which further increases with age.[3-4] Statins are proven to reduce plasma LDL-cholesterol and mortality including in the Scandinavian Simvastatin Survival Study (4S), where simvastatin caused a 35% mean reduction in plasma LDL and 30% reduction in fatal outcomes compared to placebo. [5] Statins have also proved to be effective in multiple large-scale trials.[6] This study will focus on five statins licensed in the United Kingdom: atorvastatin, fluvastatin, pravastatin, rosuvastatin and simvastatin.[7]

An adverse drug reaction (ADR) is an unintended harmful reaction to the use of a drug.[8] The degree of harm may range from a mild effect through to permanent or fatal outcomes. Statins like all drugs have their own unique ADRs: muscle related ADRs concurrent with statin prescribing such as muscle pain (myalgia) being the most reported ADR.[9]

Mechanisms have been proposed for statin-induced myopathy however there is no singular agreed pathway. A mechanism involving the dissociation of the FKBP12 binding protein, from sarcoplasmic reticulum Ca^2+^ channels in myocytes, causing pro-apoptotic signalling; but these effects were also present in patients who had not experienced myalgia so may only affect individuals susceptible due to genetics /other factors.[10-11] Higher intensity statins which cause a ≥50% reduction in LDL-cholesterol[12] have been associated with increased risk of myopathy, which also brings the pharmacokinetic parameters of the statins into consideration.[13-14]

We herein report an approach to identifying patterns between the statin structures, their unique pharmacology and suspected ADR signals.[15-19]

## Aims

The primary aim of this research is to explore suspected ADR data of atorvastatin, fluvastatin, pravastatin, rosuvastatin and simvastatin using a national registry approach. A secondary aim is to determine whether there is a link between the physicochemical and pharmacological properties of these statins and their associated side effects.

## Methods

### Chemical properties and pharmacology

The chemical database of bioactive molecules with drug-like properties, European Molecular Biology Laboratory (ChEMBL)[20] and Electronic Medicines Compendium (EMC)[21] were used to obtain the physicochemical properties and pharmacokinetic parameters for atorvastatin, fluvastatin, pravastatin, rosuvastatin and simvastatin.

Physicochemical properties included p*K*_a_, cLog_10_P, cLog_10_D_7.4_, topological polar surface area (^*t*^PSA) and the number of hydrogen bond donors (HBD) and acceptors (HBA). Certain properties increase the propensity of a molecule to penetrate the blood brain barrier (BBB)[22-23] as follows: a molecular weight of <450 Da, a neutral or basic drug molecule, the molecule not being a substrate of P-glycoprotein, <6 HBD and <2 HBA, a ^*t*^PSA of <90 Å^2^ and a log_10_D of 1-3 at p*H* = 7.4. Penetration of the BBB can lead to potential neurological side effects.

Lipophilic ligand efficiency (LLE) was calculated where LLE = pIC_50_ – cLog_10_P. pIC_50_ is the negative log_10_ of the IC_50,_ which is the concentration of drug needed to inhibit 50% of the activity of a process or response at a receptor; median IC_50_ values for each statin acting on HMG-CoA reductase were used. cLog_10_P is the calculated partition coefficient of a substance in its neutral form between an aqueous and organic phase. The LLE is a measure of the specificity of a molecule for its target accounting for its partitioning in the organic phase.[24] An LLE of >5 is associated with a significantly smaller risk of toxicity.[25]

Pharmacokinetic properties were obtained from the EMC and included the bioavailability, half-life, CYP450 activity and the degree of plasma protein binding. Experimental C_max_ values were obtained from literature databases by searching the drug name and C_max_. [26-30] The volume of distribution was obtained from the EMC, Drugbank,[31] and from a trial for simvastatin.[32] Literature searches also determined if the statins were P-glycoprotein substrates.[33]

### Pharmacological interactions

The ChEMBL database was used to identify interactions between each statin and *homosapien* proteins/targets. Median IC_50_ values were used to select a representative value from multiple laboratories. A cut-off of <10 μM was used to remove interactions that were unlikely to occur due to the inability of the statins to reach such concentrations in the body.

### Suspected Adverse Drug Reaction (ADR) Data

Suspected ADR data was obtained from the Medicines and Healthcare products Regulatory Agency’s (MHRA) Yellow Card interactive Drug Analysis Profiles (iDAPs).[34]. Suspected ADR reports from January 2018 to August 2022 were collected for each statin. This data included the number of ADRs reported to the Yellow Card scheme with reports being categorised based on the MedDRA organ classification. Organ classes of interest were identified using percentages of the total number of ADRs for that drug; an organ class was used if it had ≥10% of the total ADRs for at least one of the five statins.

### Prescribing Data

Prescribing data was obtained from OpenPrescribing Database[35] for the January 2018 to August 2022 period. Standardisation was performed by calculating the number of ADRs per 100,000 *R*_*x*_ for each statin.

### Statistical Analysis

A chi-squared analysis was done on the standardised ADRs per 100,000 *R*_*x*_ using Microsoft Excel for Mac (version 16.67) to determine the statistical significance of suspected ADR signals (*P*-value < 0.05).

### Ethical Approval

No ethical approval was required as the study used publicly available open-source data that was fully anonymised.

## Results

### Physicochemical properties and pharmacokinetics

**Table 1** shows the properties of the statins. Rosuvastatin and pravastatin were predicted to be less likely than other statins to cause toxicity based on the LLE – both being > 5; atorvastatin had the smallest LLE suggesting it may have more off-target effects compared to other statins.

**Table 1.** Physicochemical and pharmacokinetic properties of the five statin tablet formulations. Key: cLog_10_P, calculated partition coefficient; LLE, lipophilic ligand efficiency; Log_10_D_7.4_ partition coefficient at pH 7.4; MW, Molecular Weight; p*K*_a_, acid dissociation constant; ^*t*^PSA, topological polar surface area; HB, hydrogen bond; C_max_, peak serum concentration; V_d_, volume of distribution; PPB, plasma protein binding. Highlighted properties are those that increase the chance of BBB penetration.

| Property vs drug | Atorvastatin | Fluvastatin | Pravastatin | Rosuvastatin | Simvastatin |
| --- | --- | --- | --- | --- | --- |
| $c\log_{10}P$ | 5.39 | 3.83 | 1.65 | 1.92 | 4.46 |
| $pIC_{50}$ | 8.06 | 7.85 | 7.52 | 8.35 | 7.59 |
| LLE | 2.67 | 4.02 | 5.87 | 6.43 | 3.13 |
| $\log_{10}D_{7.4}$ | 2.43 | 1.05 | -1.38 | -1.24 | 4.46 |
| MW (Da) | 558.65 | 411.47 | 424.53 | 481.55 | 418.57 |
| $pK_a$ | 4.31 | 4.54 | 4.21 | 4 | Neutral |
| tPSA (Å) | 111.79 | 82.69 | 124.29 | 140.92 | 72.83 |
| HB Acceptors | 5 | 4 | 6 | 7 | 5 |
| HB Donors | 4 | 3 | 4 | 3 | 1 |
| Bioavailability (%) | 12 | 24 | 17 | 20 | <5 |
| $C_{max}$ (nM) | 118.5 | 687.78 | 189.57 | 39.04 | 130.71 |
| Half-life (h) | 14 | 2.3 | 1.5-2 | 19 | 1.9 |
| $V_d$ (L) | 381 | 330 | 0.5L/Kg | 134 | 233 |
| PPB | ≥ 98% | > 98% | 50% | 90% | > 95% |
| P-Glycoprotein substrate | Yes | No | No | No | Likely |
| Liver CYP<sub>450</sub> metabolism | 3A4 | 2C9, 3A4, 2C8 | Minimal | Minimal | 3A4, 3A5, 2C8, 2C9 |
| Dosing regime | 10-80mg OD | 20-80mg OD/BID | 10-40mg OD | 5-20mg OD | 10-80mg OD |

Fluvastatin was predicted to be the most likely to cross the BBB followed by simvastatin, pravastatin, rosuvastatin and atorvastatin based on the physicochemical properties that they possess: <450 Da MW, a tPSA < 90Å, molecule is basic or neutral, a log_10_D_7.4_ of 1-3, <2 HBA and <6 HBD and not being a substrate for P-glycoprotein transporter. Atorvastatin was found to be the most lipophilic which is reflected in its high volume of distribution (V_d_).,

### Target Affinity

**Table 2** shows the pharmacological interactions of the five statins as median IC_50_ values. Interactions with a respective IC_50_ >> C_max_ were excluded from further analysis as it is unlikely that the statins would reach these clinically relevant concentrations in the body.

**Table 2.**
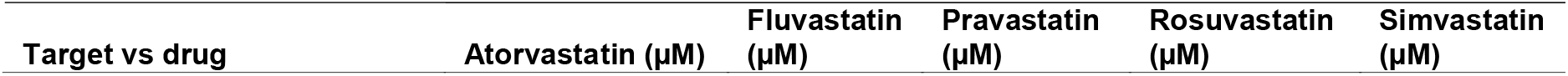
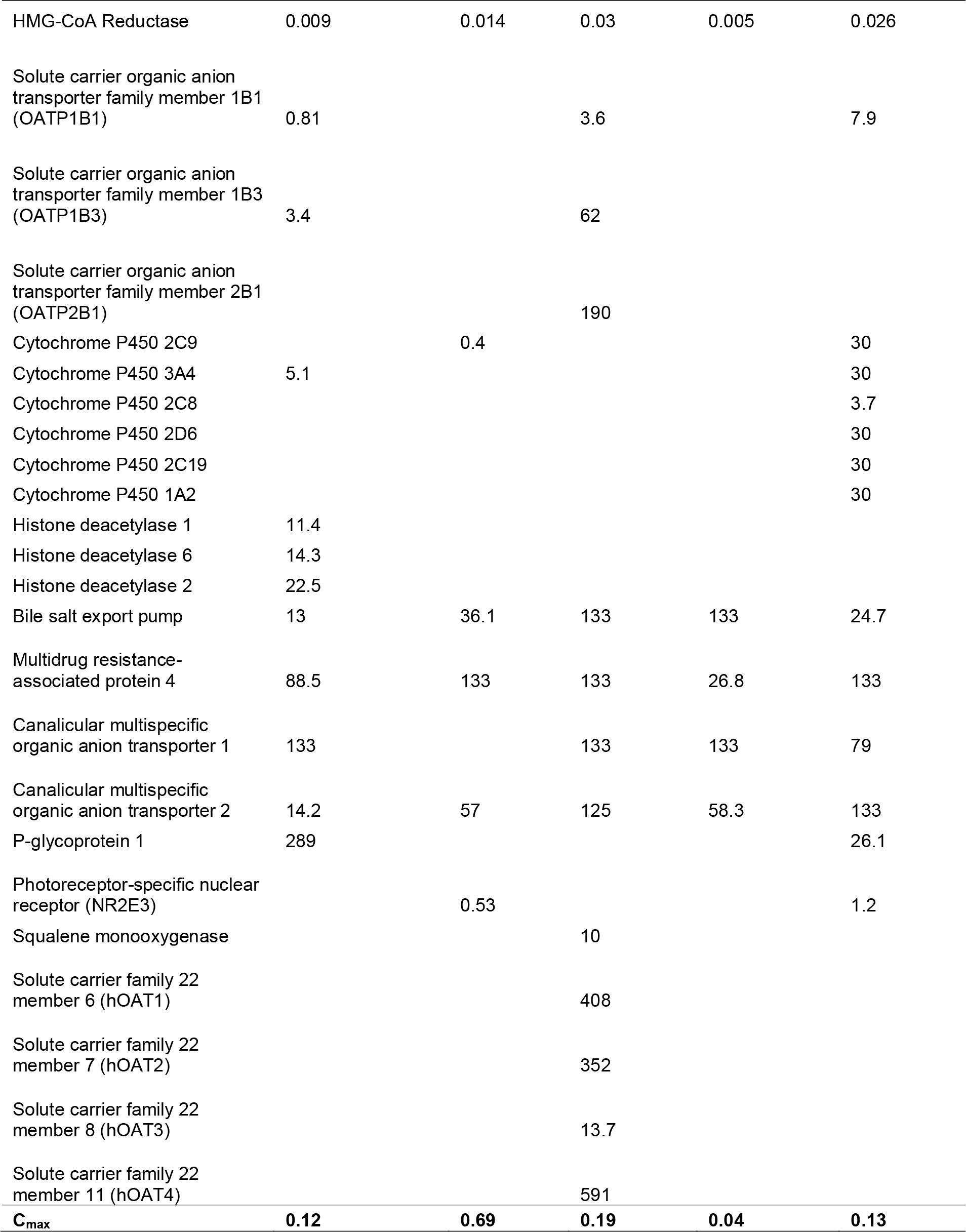
Pharmacological interactions of the statins studied.

Fluvastatin (n=2) was found to have the most potential off-target interactions that were at clinically achievable concentrations (Cytochrome P450 2C9 and Photoreceptor-specific nuclear receptor (NR2E3), respectively). Rosuvastatin, pravastatin, atorvastatin, and simvastatin had no relevant off-target interactions (n=0).

Atorvastatin, pravastatin and simvastatin showed activity at OATP1B1 with atorvastatin having the most potent action with an IC_50_ of 0.81 μM (C_max_ = 0.12 μM); atorvastatin also showed activity at OATP1B3 (3.4 μM).

Atorvastatin, fluvastatin and simvastatin showed activity at several CYP450 enzymes involved in their metabolism; CYP3A4 for atorvastatin with IC_50_ of 5.1 μM, CYP2C9 for fluvastatin with an IC_50_ of 0.4 μM and CYP2C8 for simvastatin with an IC_50_ of 3.7 μM. Fluvastatin was twice as potent as simvastatin when acting on NR2E3 (IC_50_ = 0.53 μM); and pravastatin was the only statin to cause inhibition on squalene monooxygenase with an IC_50_ of 10 μM.

### Adverse Drug Reactions

The total number of each statin prescribed in the UK, the number of suspected ADRs, and their incidence rates for the selected organ classes from January 2018 - August 2022 alongside chi-squared statistical analysis results are presented in **Table 3**.

**Table 3.**
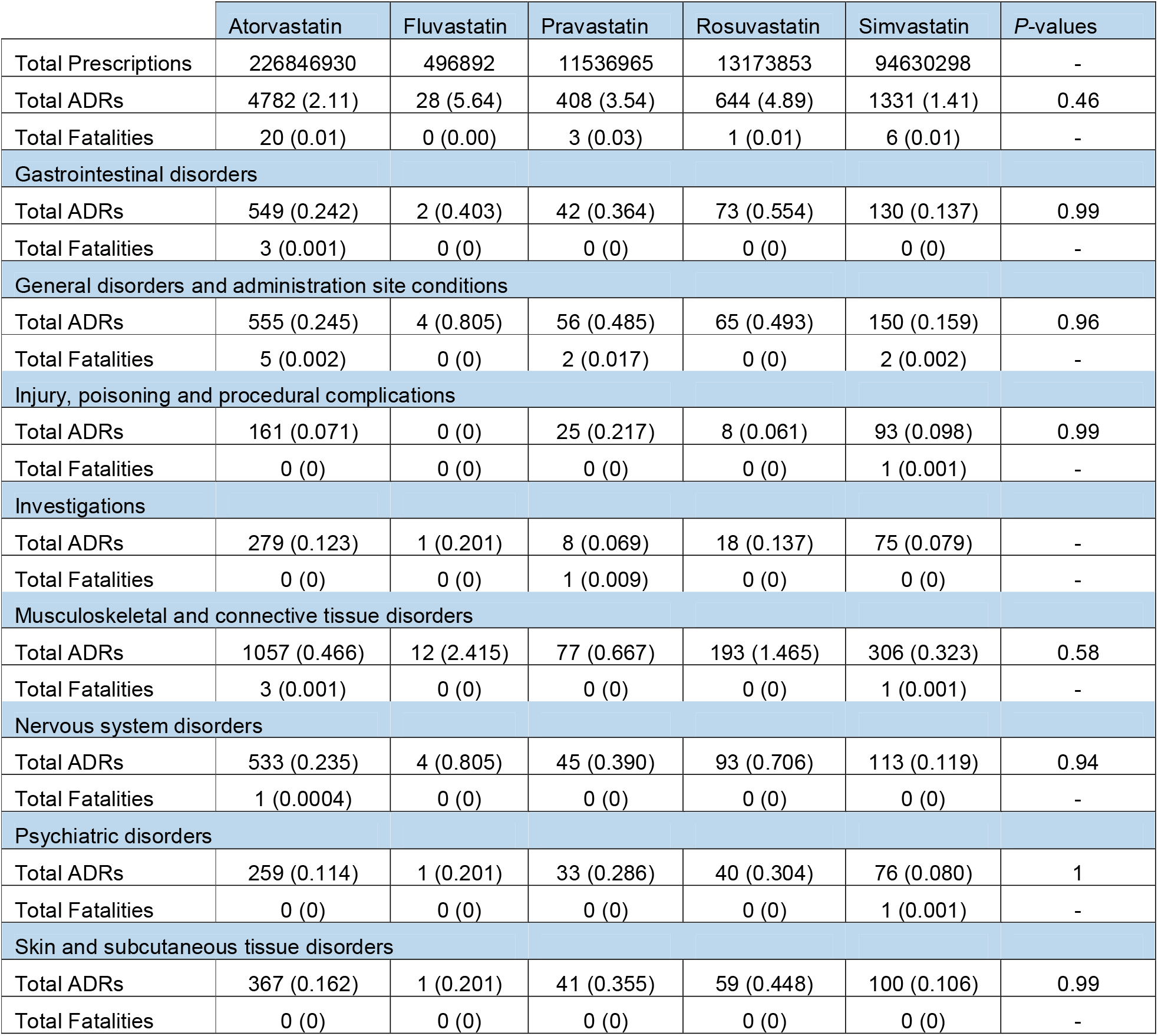
Summary of the selected Yellow Card ADR reporting data for the five statins in the UK. The numbers in the brackets are ADRs/100,000 *R*_*x*_.

Atorvastatin was the most prescribed statin (226,846,930) followed by simvastatin (94,630,298), rosuvastatin (13,173,853), pravastatin (11,536,965) and then fluvastatin with the least prescriptions (496,892).

Fluvastatin had the most reported suspected ADRs per 100,000 prescriptions (5.64) followed by rosuvastatin (4.89), pravastatin (3.54), atorvastatin (2.11) and then simvastatin (1.41). Fatality incidence was similar across the statins with rates ranging between 0.00-0.03 per 100,000 *R*_*x*_.

There was no significant difference between the suspected ADRs per 100,000 *R*_x_ for the statins (*X*^2^-analysis), in any of the organ classes however, the muscoskeletal and connective tissue class did have a noticeably different *P*-value (0.58) compared to all other organ classes.

Within the muscoskeletal and connective tissue organ class, fluvastatin had the highest incidence of ADRs (2.415) followed by rosuvastatin (1.465), pravastatin (0.667), atorvastatin (0.466) and lastly simvastatin (0.323). Atorvastatin and simvastatin had a 0.001 incidence rate per 100,000 *R*_x_ for fatalities whereas the other three statins had no reports in this time frame. Further chi-squared statisticalanalysis was performed within the muscoskeletal and connective tissue organ class (**Table 4**).

**Table 4.** Chi-squared analysis results from comparing the statins suspected ADR reports per 100,000 *R*_x_ within the muscoskeletal and connective tissue organ class.

| Statins | *P*-value |
| --- | --- |
| fluvastatin vs atorvastatin | 0.15 |
| fluvastatin vs pravastatin | 0.17 |
| fluvastatin vs rosuvastatin | 0.17 |
| fluvastatin vs simvastatin | 0.14 |
| atorvastatin vs simvastatin | 0.35 |
| atorvastatin vs pravastatin | 0.48 |
| atorvastatin vs rosuvastatin | 0.49 |
| pravastatin vs rosuvastatin | 0.59 |
| pravastatin vs simvastatin | 0.41 |
| rosuvastatin vs simvastatin | 0.41 |

There was no statistically significant difference between any pair of statins for p <0.05 however the p-values for fluvastatin were noticeably pronounced than for other statins.

## Discussion

Many of the polypharmacological interactions are unlikely to occur as the concentrations required for an effect are not clinically relevant based on the C_max_ of the statins (**Table 1**). Without the accumulation of statin, it is unlikely that plasma concentrations of statin will reach these figures. It is possible for fluvastatin to accumulate in patients with hepatic impairment[36] due to it being primarily excreted via the bile with extensive pre-systemic metabolism; this may contribute to fluvasatain having the highest incidence of ADRs per 100,000 *R*_x_ (**Table 3**). Fluvastatin had the highest ADR incidence in multiple organ classes however it was prescribed over twenty-fold less than the next least prescribed statin, pravastatin.

### ADR Incidence

Overall, no significant difference was found between the statins and any organ class for *P* < .05 (**Table 3**). This suggests that the statins could have a similar class effect not individual differing off-target pharmacological mechanisms.

For the muscoskeletal and connective tissue organ class (*P* = .58) there was no statistically significant difference between the statins however, when compared to the *P* values for other organ classes - a difference in risk emerges. Further analysis within the muscoskeletal and connective tissue organ class showed no statistically significant difference (**Table 4**) however, fluvastatin compared to other statin-pairs had lower *P* valies and also had the highest suspected ADR incidence across multiple organ classes – which was unexpected.

Fluvastatin is a low-medium intensity statin.[37] The National Institute for Health and Care Excellence (NICE) guidelines, recommend a high intensity statin as first line treatment for patients at risk of cardiovascular disease.[38] Another contributing factor is that fluvastatin is the only statin indicated for use after percutaneous coronary intervention, a procedure which is also known to cause some pain post-operatively.[39] The low prescribing rate, potential for accumulation and patient-comorbidities could explain the unexpectedly high suspected ADR incidence of fluvastatin.

Chi-squared analysis has shown that there was no significant difference between the statins in the muscoskeletal and connective tissue organ class and amongst the remaining ADR organ classes. These findings confirm a recent meta-analysis[14] that there was no clear evidence that the risk ratios for muscoskeletal symptoms differed between statins; it did however find that higher intensity statins caused an increased risk of muscle pain or weakness compared to moderate intensity statins. There is no sequential change of intensity between the statins when looking at the suspected muscle ADR incidence (**Table 3**); Rosuvastatin is classed as moderate-high statin which had the next highest suspected muscle ADR incidence followed by pravastatin which is classed as low-moderate statin.

These findings do also align with another large-scale meta-analysis[40] which found that patients were less likely to experience myalgia with simvastatin than with atorvastatin in a pairwise meta-analysis; it also found no significant difference between the statins when collectively comparing 1,986 myalgia events in a drug-level network meta-analysis.

### Physicochemical properties

There does not appear to be a clear relationship between the physicochemical properties (**Table 1**) and the suspected ADR incidence. Rosuvastatin and pravastatin are the most hydrophilic compared to the remaining statins based on their negative log_10_D^7.4^ values. This prevents them from passively diffusing through tissue and requires the use of carriers to facilitate their uptake into the liver.[41] This should in theory increase selectivity and so reduce uptake into other tissues such as muscle tissue however, this is not reflected in the suspected muscle ADR incidence as the lipophilic atorvastatin and simvastatin had smaller suspected incidence values compared to the two hydrophilic statins (rosuvastatin and pravastatin).

Based on the physicochemical properties, fluvastatin was predicted as most likely to cross the BBB followed by simvastatin, pravastatin, rosuvastatin and atorvastatin. Previous studies have found that the lipophilic statins such as atorvastatin, fluvastatin and simvastatin can easily cross the BBB whilst hydrophilic statins are less likely to achieve this at clinically used concentrations. [42] There does not appear to be a relationship between the statins which were predicted as more likely to cross the BBB and ADR incidence in the nervous system and psychiatric disorder categories. It is unclear based on current research whether there are any causative links between statins and these types of disorders or whether some individuals are predisposed to these conditions and are affected by them coincidentally during their treatment with statins.[43]

### Pharmacological interactions

Atorvastatin, pravastatin, and simvastatin showed modest activity at OATP1B1 transporter whilst atorvastatin also showed activity at OATP1B3. Statins are known to be substrates of these organic anion transporter polypeptides which facilitate uptake into the liver.[44] Mutations in the SLCO1B1 gene which encodes for OATP1B1 have been linked to decreased hepatic uptake of statins and increased systemic exposure,[45] increasing the risk of myopathy. Patients taking inhibitors of these transporters such as ciclosporin[46] are advised to reduce their statin dose to prevent ADRs because of increased statin exposure.

Fluvastatin showed activity at CYP2C9, atorvastatin at CYP3A4 and simvastatin at CYP2C8. These are enzymes that are involved in the metabolism of these statins and unlikely to have a role in ADRs unless the statins are taken concomitantly with inhibitors of these enzymes, in turn increasing systemic exposure to the statin. Inhibitors of cytochrome P450 enzymes are less likely to affect rosuvastatin and pravastatin as these are metabolised via other pathways.[41]

Pravastatin showed inhibition of squalene monooxygenase which is another rate-limiting enzyme in the cholesterol synthesis pathway acting downstream of HMG-CoA reductase.[47] Little evidence is available for the clinical relevance of squalene monooxygenase inhibition, with animal studies showing symptoms of dermatitis and neuropathy due to squalene monooxygenase inhibitors.[48]

Fluvastatin and simvastatin showed activity at the photoreceptor-specific nuclear receptor (NR2E3) which is involved in photoreceptor proliferation. Mutations of its gene have been shown to cause retinal degeneration in animal studies and other eye disorders.[49] It is unclear whether this interaction has any significance for statins and in causing ADRs, no studies of clinical relevance regarding this interaction were identified.

## Limitations

Prescribing data was available from November 2017 until August 2022 however due to the format in which the ADR data was available from the MHRA Yellow card scheme, data was extracted from January 2018 (excluding the data available from 2017). However, this ensured that the suspected ADR incidence per 100,000 *R*_*x*_ for 2017 was not inflated due to ADRs being included for the two months in 2017 where prescribing data was not available.

Reports from the Yellow Card Scheme are suspected reports and therefore no causal relationship must be demonstrated or evidenced before submitting a report. Therefore, reported ADRs in any registry may have no defined relationship to the pharmacology of the statins. Comparison of the ADR incidence for a statin against an average incidence was used to ensure relevant ADRs were highlighted. Underreporting of ADRs is also a common issue with the pharmacovigiance schemes leading to ADRs going undetected.[50] Information such as other drug use and health conditions are not available from the Yellow Card scheme and so it is not possible to establish causality through this data alone.

The muscoskeletal and connective tissue disorders organ class was analysed as a whole and so included connective tissue disorders in the statistical analysis. Interactions with human proteins were extracted from the ChEMBL database which necessarily does not contain every possible interaction which the statins could have in the human body. Furthermore, multiple IC_50_ values were available for each protein and so a median value was used.

## Conclusions

Statins have proven to be effective in the reduction of cardiovascular events with a small risk of muscle related ADRs that is outweighed by the benefits to the patient. The study found that there was no significant difference between the statins across the organ classes investigated and specifically in the muscoskeletal and connective tissue disorder category. Fluvastatin was found to have an unexpectedly high suspected ADR incidence across multiple organ classes which initially could be attributed to lower prescribing rates than the other statins but was corrected for in this study based on prescribing levels.

Pharmacological interactions of the statins included the cytochrome P450 enzymes and the organic anion transporters. New interactions with NR2E3 and squalene monooxygenase were identified. However, a relationship to statin ADRs was not clear.

## Data Availability

All data produced in the present work are contained in the manuscript and supporting information.

## Conflict of Interest

There is no conflict of interest to be disclosed.

## Availability of Data

The data that support the findings of this study are openly available in the Supporting Information file.

## Notes

### Competing Interest Statement

The authors have declared no competing interest.

### Funding Statement

This study did not receive any funding.

### Author Declarations

The study used ONLY openly available human data that were originally located at: https://yellowcard.mhra.gov.uk/idaps

