## Supplementary Information for "The Relationship Between Suspected Adverse Drug Reactions Of HMG-Coa Reductase Inhibitors And Polypharmacology Using A National Registry Approach"

**CONTENTS**

| Chart S1 | Page 2 |
| --- | --- |
| Table S2 | Page 3 |
| Table S3 | Page 5 |
| Chart S4 | Page 7 |

**Chart S1.** The number of prescriptions dispensed per statin between January 2018 and August 2022. (Fluvastatin; 496,892)

Number of items (Total Prescriptions)

|  | Statins |
| --- | --- |
| Atorvastatin | 226846930 |
| Fluvastatin | 496892 |
| Pravastatin | 11536965 |
| Rosuvastatin | 13173853 |
| Simvastatin | 94630298 |

**Table S2.** Non-fatal ADRs per 100,000 prescriptions of the statins and chi-squared results for all organ classes (selected organ classes are highlighted)

|  | Atorvastatin | Fluvastatin | Pravastatin | Rosuvastatin | Simvastatin | X^2^ |
| --- | --- | --- | --- | --- | --- | --- |
| Blood and lymphatic system disorders | 0.015 | 0.000 | 0.026 | 0.000 | 0.012 | 1.00 |
| Cardiac disorders | 0.027 | 0.201 | 0.052 | 0.046 | 0.023 | 0.99 |
| Congenital, familial and genetic disorders | 0.000 | 0.000 | 0.000 | 0.008 | 0.007 | 1.00 |
| Ear and labyrinth disorders | 0.020 | 0.000 | 0.009 | 0.023 | 0.002 | 1.00 |
| Endocrine disorders | 0.002 | 0.000 | 0.000 | 0.000 | 0.000 | 1.00 |
| Eye disorders | 0.038 | 0.000 | 0.069 | 0.068 | 0.029 | 1.00 |
| Gastrointestinal disorders | 0.242 | 0.403 | 0.364 | 0.554 | 0.137 | 0.99 |
| General disorders and administration site conditions | 0.245 | 0.805 | 0.485 | 0.493 | 0.159 | 0.96 |
| Hepatobiliary disorders | 0.046 | 0.000 | 0.026 | 0.000 | 0.015 | 1.00 |
| Immune system disorders | 0.011 | 0.000 | 0.026 | 0.046 | 0.016 | 1.00 |
| Infections and infestations | 0.029 | 0.000 | 0.069 | 0.023 | 0.025 | 1.00 |
| Injury, poisoning and procedural complications | 0.071 | 0.000 | 0.217 | 0.061 | 0.098 | 0.99 |
| Investigations | 0.123 | 0.201 | 0.069 | 0.137 | 0.079 | 1.00 |
| Metabolism and nutrition disorders | 0.034 | 0.000 | 0.121 | 0.083 | 0.034 | 1.00 |
| Musculoskeletal and connective tissue disorders | 0.466 | 2.415 | 0.667 | 1.465 | 0.323 | 0.58 |
| Neoplasms benign, malignant and unspecified (incl cysts and polyps) | 0.005 | 0.000 | 0.000 | 0.000 | 0.007 | 1.00 |
| Nervous system disorders | 0.235 | 0.805 | 0.390 | 0.706 | 0.119 | 0.94 |
| Pregnancy, puerperium and perinatal conditions | 0.001 | 0.000 | 0.000 | 0.000 | 0.000 | 1.00 |
| Product issues | 0.027 | 0.201 | 0.009 | 0.091 | 0.008 | 0.98 |
| Psychiatric disorders | 0.114 | 0.201 | 0.286 | 0.304 | 0.080 | 1.00 |
| Renal and urinary disorders | 0.050 | 0.201 | 0.087 | 0.083 | 0.049 | 1.00 |
| Reproductive system and breast disorders | 0.025 | 0.000 | 0.000 | 0.046 | 0.003 | 1.00 |
| Respiratory, thoracic and mediastinal disorders | 0.091 | 0.000 | 0.147 | 0.159 | 0.052 | 1.00 |
| Skin and subcutaneous tissue disorders | 0.162 | 0.201 | 0.355 | 0.448 | 0.106 | 0.99 |
| Social circumstances | 0.003 | 0.000 | 0.009 | 0.008 | 0.003 | 1.00 |
| Surgical and medical procedures | 0.000 | 0.000 | 0.009 | 0.008 | 0.010 | 1.00 |
| Vascular disorders | 0.025 | 0.000 | 0.043 | 0.030 | 0.010 | 1.00 |
| Total Reactions | 2.11 | 5.64 | 3.54 | 4.89 | 1.41 | 0.46 |

**Table S3.** Fatal ADRs per 100,000 prescriptions of the statins and chi-squared results for all organ classes (selected organ classes are highlighted)

|  | Atorvastatin | Fluvastatin | Pravastatin | Rosuvastatin | Simvastatin | X^2^ |
| --- | --- | --- | --- | --- | --- | --- |
| Blood and lymphatic system disorders | 0.015 | 0.000 | 0.026 | 0.000 | 0.012 | 1.00 |
| Cardiac disorders | 0.027 | 0.201 | 0.052 | 0.046 | 0.023 | 0.99 |
| Congenital, familial and genetic disorders | 0.000 | 0.000 | 0.000 | 0.008 | 0.007 | 1.00 |
| Ear and labyrinth disorders | 0.020 | 0.000 | 0.009 | 0.023 | 0.002 | 1.00 |
| Endocrine disorders | 0.002 | 0.000 | 0.000 | 0.000 | 0.000 | 1.00 |
| Eye disorders | 0.038 | 0.000 | 0.069 | 0.068 | 0.029 | 1.00 |
| Gastrointestinal disorders | 0.001 | 0.000 | 0.000 | 0.000 | 0.000 | 1.00 |
| General disorders and administration site conditions | 0.002 | 0.000 | 0.017 | 0.000 | 0.002 | 1.00 |
| Hepatobiliary disorders | 0.046 | 0.000 | 0.026 | 0.000 | 0.015 | 1.00 |
| Immune system disorders | 0.011 | 0.000 | 0.026 | 0.046 | 0.016 | 1.00 |
| Infections and infestations | 0.029 | 0.000 | 0.069 | 0.023 | 0.025 | 1.00 |
| Injury, poisoning and procedural complications | 0.000 | 0.000 | 0.000 | 0.000 | 0.001 | 1.00 |
| Investigations | 0.000 | 0.000 | 0.009 | 0.000 | 0.000 | 1.00 |
| Metabolism and nutrition disorders | 0.034 | 0.000 | 0.121 | 0.083 | 0.034 | 1.00 |
| Musculoskeletal and connective tissue disorders | 0.001 | 0.000 | 0.000 | 0.000 | 0.001 | 1.00 |
| Neoplasms benign, malignant and unspecified (incl cysts and polyps) | 0.005 | 0.000 | 0.000 | 0.000 | 0.007 | 1.00 |
| Nervous system disorders | 0.000 | 0.000 | 0.000 | 0.000 | 0.000 | 1.00 |
| Pregnancy, puerperium and perinatal conditions | 0.001 | 0.000 | 0.000 | 0.000 | 0.000 | 1.00 |
| Product issues | 0.027 | 0.201 | 0.009 | 0.091 | 0.008 | 0.98 |
| Psychiatric disorders | 0.000 | 0.000 | 0.000 | 0.000 | 0.001 | 1.00 |
| Renal and urinary disorders | 0.050 | 0.201 | 0.087 | 0.083 | 0.049 | 1.00 |
| Reproductive system and breast disorders | 0.025 | 0.000 | 0.000 | 0.046 | 0.003 | 1.00 |
| Respiratory, thoracic and mediastinal disorders | 0.091 | 0.000 | 0.147 | 0.159 | 0.052 | 1.00 |
| Skin and subcutaneous tissue disorders | 0.000 | 0.000 | 0.000 | 0.000 | 0.000 | - |
| Social circumstances | 0.003 | 0.000 | 0.009 | 0.008 | 0.003 | 1.00 |
| Surgical and medical procedures | 0.000 | 0.000 | 0.009 | 0.008 | 0.010 | 1.00 |
| Vascular disorders | 0.025 | 0.000 | 0.043 | 0.030 | 0.010 | 1.00 |

**Chart S4.** The total number of suspected ADRs per 100,000 prescriptions for the statins between 2018 and 2022 for the selected organ classes.

Total number of ADRs per 100,000 prescriptions

| Organ Class | Atorvastatin | Fluvastatin | Pravastatin | Rosuvastatin | Simvastatin |
| --- | --- | --- | --- | --- | --- |
| Gastrointestinal disorders | 0.242013 | 0.402502 | 0.364047 | 0.554128 | 0.137377 |
| General disorders and administration site conditions | 0.244658 | 0.805004 | 0.485396 | 0.493402 | 0.158512 |
| Injury, poisoning and procedural complications | 0.070973 | 0 | 0.216695 | 0.0607263 | 0.0982772 |
| Investigations | 0.12299 | 0.201251 | 0.0693423 | 0.136634 | 0.0792558 |
| Musculoskeletal and connective tissue disorders | 0.465953 | 2.41501 | 0.66742 | 1.46502 | 0.323364 |
| Nervous system disorders | 0.23496 | 0.805004 | 0.390051 | 0.705944 | 0.119412 |
| Psychiatric disorders | 0.114174 | 0.201251 | 0.286037 | 0.303632 | 0.0803125 |
| Skin and subcutaneous tissue disorders | 0.161783 | 0.201251 | 0.355379 | 0.447857 | 0.105674 |
